# Kinetical analysis of D-alanine upon oral intake in humans

**DOI:** 10.1101/2024.05.21.24307633

**Authors:** Tomonori Kimura, Shinsuke Sakai, Masaru Horio, Shiro Takahara, Shoto Ishigo, Maiko Nakane, Eiichi Negishi, Hiroshi Imoto, Masashi Mita, Kenji Hamase, Yoko Higa-Maekawa, Yoichi Kakuta, Masayuki Mizui, Yoshitaka Isaka

## Abstract

D-Alanine, a rare enantiomer of alanine in life, can potentially alleviate the worsening of viral infections and maintain circadian rhythm. This study aimed to analyze the kinetics of D-alanine upon oral intake. Five healthy volunteers were administered D-alanine as a single oral dose at 12,366 or 33,708 μmoL. Upon intake of the lower dose, the plasma level of D-alanine reached its peak concentration of 588.4 ± 40.9 μM with a peak time of 0.60 ± 0.06 h. The plasma level of D-alanine became close to the endogenous level after 24 h. The compartment model estimated the clearance of D-alanine at 12.5 ± 0.3 L/h, or 208 ± 5 mL/min, distribution volume of 8.3 ± 0.7 L and half-life of 0.46 ± 0.04 h. The peak concentration and area under the curve increased proportionally upon intake of the higher dose, while the clearance, distribution volume and half-life did not. The urinary ratio of D-alanine reached its peak of nearly 100%, followed by a slow decline. The peak time of the urinary ratio was 1.15 ± 0.15 h, showing a time lag of blood to urine excretion. Fractional excretion of D-alanine increased from 14.0 ± 5.8% to 64.5 ± 10.3%; the latter corresponded to the urinary clearance of D-alanine as about 77 mL/min for an adult, with a peak time of 1.90 ± 0.56 h. D-Alanine was quickly absorbed and appeared in blood, followed by urinary excretion. This kinetic analysis increases our fundamental knowledge of the oral intake of D-alanine.

## Introduction

D-Alanine is a natural nutrient. D-Amino acids, including D-alanine, are the enantiomers of L-amino acids that are dominant in life. Recent studies revealed that D-alanine is present in mammals, including humans (1-3), and is connected to many physiological functions and diseases (4). The blood level of D-alanine decreases in severe viral infections such as COVID-19 or influenza virus infection (5, 6), while it increases in patients with kidney diseases (3, 7). D-Alanine can serve as a co-agonist of the *N*-methyl-D-aspartate (NMDA) receptor (8) and activate macrophages (9). The protective effect of D-alanine against diseases has also been demonstrated in rodent models. These include viral infections such as COVID-19 and influenza viral infection (5), experimental colitis (10) and acute kidney injury (11). D-Alanine also has a close association with the circadian rhythm (12). Meanwhile, D-alanine has a clear circadian rhythm (13) and maintains this rhythm in a rodent model (12). Through the regulation of circadian rhythm, D-alanine maintains physical conditions, such as sleep and activity, glucose production, and potentially immune responses (12).

A small proportion of alanine in blood is D-alanine, while the rest is L-alanine (3, 7). D-Alanine is not synthesized in mammalian cells. Therefore, D-alanine in mammals is of external origins, such as food and the intestinal microbiome (14). D-Alanine is relatively rich in fermented foods or fish from brackish water (15-18). D-Alanine in mice is also derived from the intestinal microbiome (14, 19). D- and L-alanine taste different, with the former tasting sweeter (20). D,L-Alanine, a form that includes both L- and D-enantiomer, has been approved as a food additive in the world including Japan and United States, and is primarily used as a seasoning (21); however, there is no food whose content of D-alanine is certificated.

Normalization of the circadian cycle by D-alanine may be therapeutic for many life style-related diseases; however, much is still not known about its mechanism of action. This includes the kinetics of D-alanine in the body (4). After oral intake and intestinal absorption, D-alanine enters the blood and is delivered to the tissues. About 20% of cardiac output is delivered to the kidney, where the blood is subjected to glomerular filtration. After filtration, a fraction of D-alanine is reabsorbed at the proximal tubules, whereas the rest is excreted into the urine. The excretion ratio was estimated to about 20% using creatinine as a reference (7, 12). The reabsorbed fraction of D-alanine is oxidized by D-amino acid oxidase (DAO), which is predominantly present in the proximal tubules (22-24), and this reaction results in the production of peroxide and pyruvate (12). The unoxidized fraction of D-alanine is thought to re-enter the bloodstream (4). D-Alanine in the blood is delivered to several tissues. Thus far, it has been detected in endocrine tissue, such as the pancreatic islets, the adrenal glands, and the pituitary gland (25), as well as the brain, liver and kidney (26).

Urinary excretion and oxidation by DAO contribute to maintaining blood D-alanine levels. The level of D-alanine drastically increases in patients with kidney diseases (3, 4, 7, 27) or *Dao*-deficient rodents (19, 26). However, the quantitative effects of these regulations are still unknown (4). D-Alanine is not incorporated in proteins by translation, and other biological processes that clear D-alanine have not been found. Therefore, this study aimed to analyze the basic dynamics of D-alanine in the body, using general kinetical analysis, and estimated the contribution of the kidney and DAO to the clearance of D-alanine.

## Methods

### Study design and participants

This is an open, non-randomized study. Participants were healthy adults aged ≥ 20 or older who took no medication in the previous month. We recruited 5 volunteers from three centers in Japan between July 2023 and December 2023. Subjects fasted after their evening meal for 10 hours. At 8 am, each received a packaged powder of D-alanine (Direct Alanine^@^, KAGAMI INC) with 200 mL of water. Blood samples (100 μL) were taken before administration, and 0.25, 0.5, 0.75, 1, 2, 4, 8, and 24 hours thereafter. Urine samples were taken after each blood sampling. The test was conducted with different volumes (1 g [11,236 μmoL] or 3 g [33,708 μmoL] of D-alanine), and each study was conducted with an interval of at least one week. Followings are the specification of Direct Alanine^@^ used in this study: amount, 2.0 ± 0.10 g per stick; alanine content, 99.0–103%; d-alanine composition ratio, 49.0–51.0%. Upon sampling inspection, the amount was measured using electron balance whereas alanine content and D-alanine composition ratio were quantified using two-dimensional high-performance liquid chromatography (2D-HPLC) as follows. This study was conducted in compliance with the Declaration of Helsinki, the Ethical Guidelines for Medical Research Involving Human Subjects. This study was registered in UMIN-CTR (#UMIN000050865). Approval for all facilities was obtained from the Central Ethics Review Committee of Osaka University (#122472). Written informed consent was obtained from all the participants.

### Factional excretion (FE)

The fractional excretion was calculated as follows: Ux (mg/dL) × Sy (mg/dL)/Sx (mg/dL) × Uy (mg/dL), where x is a substrate substance, y is a reference substance, Ux is the urinary concentration of x, Sy is the plasma concentration of y, Sx is the plasma concentration of x, and Uy is the urinary concentration of y. As a reference, we used D-asparagine and D-serine based on close dynamics to inulin, a gold standard for measuring kidney function (28, 29).

### Quantification of D-amino acids

Sample preparations and quantification of amino acid enantiomers by a 2D-HPLC system were performed as previously described (30, 31). Briefly, twenty-fold volumes of methanol were added to the sample and an aliquot (10 μL of the supernatant obtained from the methanol homogenate) was placed in a brown tube. After drying the solution under reduced pressure, 20 μL of 200 mM sodium borate buffer (pH 8.0) and 5 μL of fluorescence labeling reagent (40 mM 4-fluoro-7-nitro-2,1,3-benzoxadiazole in anhydrous acetonitrile) were added and then heated at 60 °C for 2 min. An aqueous solution of 0.1% (v/v) trifluoroacetic acid (75 μL) was added, and 2 μL of the reaction mixture was subjected to the 2D-HPLC.

The enantiomers of the amino acids were quantified using the 2D-HPLC platform. The fluorescence-labeled amino acids were separated using a reversed-phase column (Singularity RP column, 1.0 mm i.d. × 50 mm; provided by KAGAMI Inc., Osaka, Japan), with the gradient elution using aqueous mobile phases containing acetonitrile and formic acid. To determine D- and L-amino acids separately, the fractions of amino acids were automatically collected using a multi-loop valve and transferred to the enantioselective column (Singularity CSP-001S, 1.5 mm i.d. × 75 mm; KAGAMI Inc.). The mobile phases are the mixed solution of methanol-acetonitrile containing formic acid, and the fluorescence detection was carried out at 530 nm with excitation at 470 nm using two photomultiplier tubes.

Target peaks were quantified by scaling the standard peak shape.^19^ In this method, the shape of a peak was used for the identification of the substrate, whereas the magnitude of the intensity was used for quantification. From the chromatogram of a sample, target shapes of amino acid enantiomers were identified based on the elution time and shape of the peak. The peak shape obtained by the standard amino acid enantiomer was superimposed to the obtained peak sections, and the magnification constant best fitted to the target peak was identified. The concentration of the target enantiomer was calculated by using an identified magnification constant and the calibration lines. The peak shape method potentiated quantification within a few seconds. The fully-automatic 2D-HPLC system required < 10 min for the measurements of each D-amino acid, including separation, identification, and quantification steps. The D-amino acid ratio was defined as the percentage (%) of D-amino acids to the sum of L- and D-amino acids.

### Kinetic study

Time courses of D-alanine plasma concentrations were analyzed by noncompartmental and compartmental methods. The area under the curve from zero to the end of the observation period was estimated using the log-linear trapezoidal rule. The selection of the end of the observation, not infinite, is based on the following: (i) the final plasma level of D-alanine was sufficiently lower than its peak, and (ii) despite the low concentration, the presence of endogenous D-alanine contradicts the hypothesis of an infinite model that the level of D-alanine regresses to zero after infinite time.

Estimation of compartmental parameters were performed for each individual after fitting the data to one- or two-compartment models. The discrimination of the models was based on the goodness-of-fit comparisons using the CV of parameter estimates and Akaike’s Information Criterion. The simple unweighted results are shown after observing no improvement in the various weighting schemes. Analyses were performed using WinNonlin Professional (version 8.0) software (Pharsight, Cary, NC).

### Statistical analysis

Data were expressed as mean ± standard deviation, or as count and ratio (%). Data visualization were performed using GraphPad Prism 8.0.

## Results

### Kinetics of D-alanine in blood

Five healthy volunteers were enrolled. After oral intake of D-alanine, either at the dose of 12,366 (low dose) or 33,708 μmoL (high dose), blood samples were collected at specific time intervals. There were no adverse events.

After oral intake of D-alanine, the level of D-alanine in the plasma quickly reached its peak concentration (Figure 1, Supplementary Figures S1 and S2). The kinetic parameters are shown in Table 1. The peak concentration (C_*max*_) was 588.4 ± 40.9 μM for the low dose and 1692.0 ± 69.3 μM for the high dose, revealing a clear proportionality with the dose. The t_*max*_ was 0.60 ± 0.06 h for low dose and 0.85 ± 0.06 h for high dose. The elimination phase began immediately after the peak, and the slope of the concentration curve was exponential. After 24 h, the plasma level of D-alanine became closer to the endogenous level. Noncompartmental modeling calculated the AUC for 24 h of 1,649 ± 58 h x μmoL/L for low dose and 4,942 ± 209 h x μmoL/L for high dose, again showing a clear dose-dependent proportionality. Regarding the analysis of L-alanine, the level of L-alanine in plasma quickly reached its peak concentration, followed by a sharp decline to a level similar to the baseline, and was relatively constant thereafter (Supplementary Figures S1 and S2). The peak levels of L-alanine in blood were similar to those of D-alanine, potentially because the powder used in this study contains identical amount of D- and L-alanine. The ratio of D-alanine in plasma, as calculated as the concentration of D-alanine per sum of L- and D-alanine, showed a similar kinetic curve as that of D-alanine (Supplementary Figures S1 and S2).

**Table 1.** Kinetic parameters of plasma D-alanine after oral intake.

| Dose | $C_0$ | $T_{max}$ | $C_{max}$ | $AUC_{last}$ | Clearance | | Distribution | Half life |
| --- | --- | --- | --- | --- | --- | --- | --- | --- |
| ($\mu\text{mol}$) | ($\mu\text{mol/L}$) | (h) | ($\mu\text{mol/L}$) | (h x $\mu\text{mol/L}$) | (L/h) | (mL/min) | volume (L) | (h) |
| 11,236 | 2.0 ± 0.4 | 0.60 ± 0.06 | 588.4 ± 40.9 | 1649.0 ± 58.0 | 12.5 ± 0.3 | 208 ± 5 | 8.3 ± 0.7 | 0.46 ± 0.04 |
| 33,708 | 1.3 ± 0.3 | 0.85 ± 0.06 | 1692.0 ± 69.3 | 4942.9 ± 208.9 | 10.5 ± 0.8 | 175 ± 14 | 8.9 ± 0.4 | 0.60 ± 0.06 |

AUC, area under curve.

**Figure 1.**
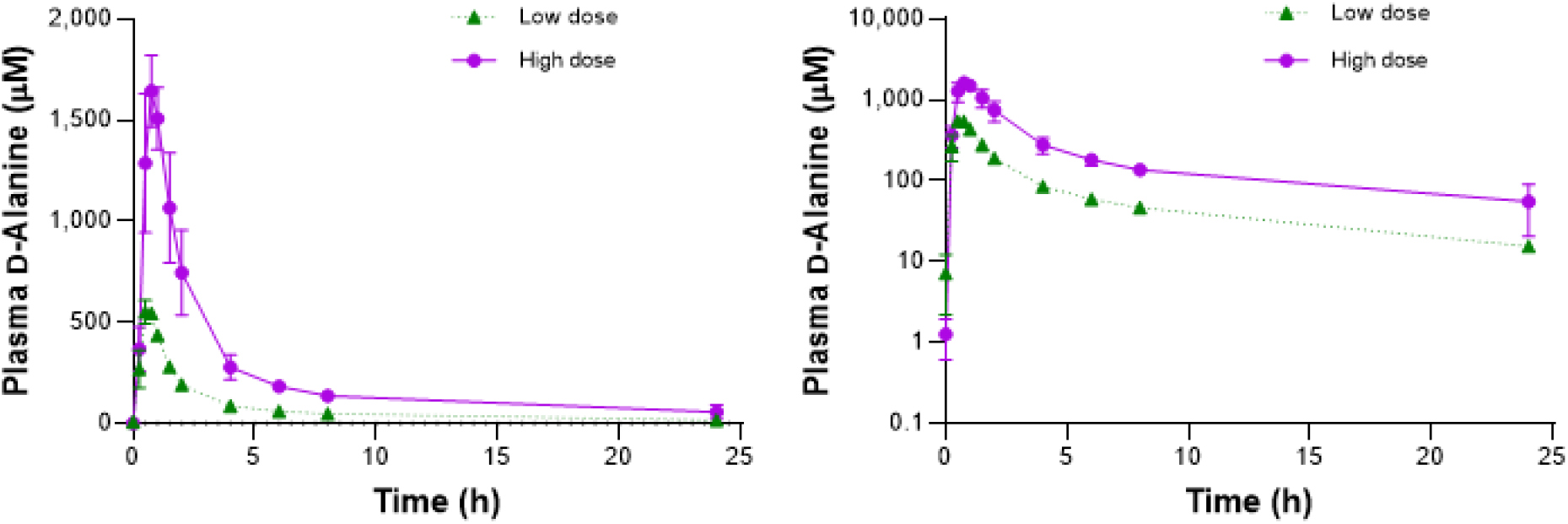
Plasma D-alanine levels after oral intake of D-alanine. Y axes with linear (left) or log-scaled (right) are shown.

Despite the low concentration, endogenously present D-alanine contradicts the hypothesis of the infinite model that the level of D-alanine regresses to zero after infinite time. Therefore, we abandoned further analysis using a non-compartment model and applied compartment models to estimate further kinetic parameters. One compartment model was selected based on the goodness-of-fit comparisons. In a small dose study, the clearance of D-alanine was estimated to be 12.5 ± 0.3 L/h, or 208 ± 5 mL/min, the distribution volume as 8.3 ± 0.7 L, and half-life as 0.46 ± 0.04 h (Table 1). The analysis for the high dose showed similar estimates (Table 1).

### Urinary kinetics of D-alanine

Since the level of D-alanine in plasma is regulated through urinary excretion, we chased the urinary level of D-alanine. Since the urinary level is subject to condensation, we calculated the urinary ratio of D-alanine per sum of D- and L-alanine for the analysis. The urinary ratio of D-alanine was initially 12.0 ± 2.4% and quickly reached nearly 100%, followed by a slow decline (Figure 2 and Table 2). The analysis of the high dose showed a similar trend (Figure 2 and Table 2). The urinary level of D-alanine quickly peaked, followed by a sharp decline (Supplementary Figures S3 and S4). The T_*max*_ was 1.15 ± 0.15 h. The plasma D-alanine level peaked earlier than the urinary level, since D-alanine enters plasma after oral intake and then enters the urine. The wide error bars of urinary levels likely reflected the variations of urinary condensation between samples. The peak urinary level of L-alanine was much lower than that of D-alanine, since almost 99% of L-alanine is reabsorbed in the kidney tubules.

**Table 2.** Urinary excretion kinetics of D-alanine.

| **Dose** | **C<sub>0</sub>** |  |  | **T<sub>max</sub>** |  |  | **C<sub>max</sub>** |  |  |
| --- | --- | --- | --- | --- | --- | --- | --- | --- | --- |
| **(μmol)** | **(%)** |  |  | **(h)** |  |  | **(%)** |  |  |
| 11,236 | 12.0 | ± | 2.4 | 1.15 | ± | 0.15 | 98.2 | ± | 0.3 |
| 33,708 | 13.9 | ± | 3.3 | 1.10 | ± | 0.10 | 99.5 | ± | 0.1 |

%, D-alanine ratio.

**Figure 2.**
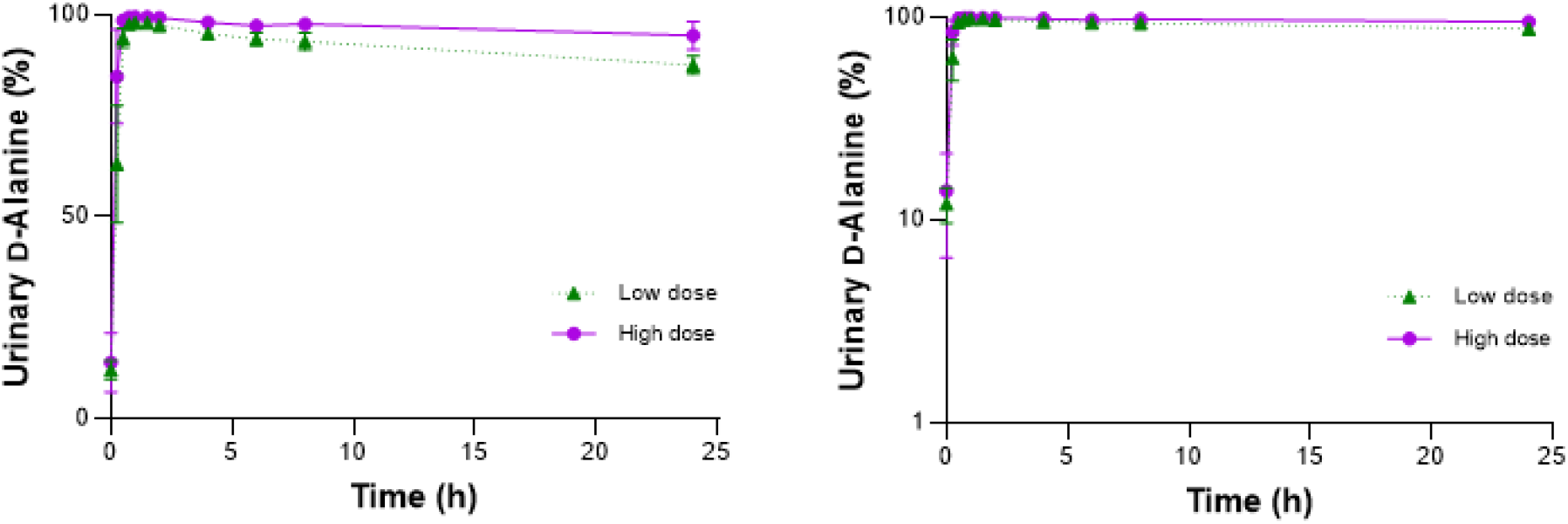
Urinary D-alanine levels or ratios after oral intake of D-alanine. Y axes with linear (left) or log-scaled (right) are shown.

To further delineate the urinary excretion dynamics, we calculated the fractional excretion (FE). FE is the ratio of the clearance of a substance to the clearance of a standard molecule. It is used to monitor the excretion ratio of the substance after glomerular filtration. FE is usually calculated using creatinine as a reference; however, this calculation could lead to an underestimation of FE since creatinine is positively excreted into urine. To avoid this, we used D-asparagine or D-serine as references, as the urinary excretion dynamics are close to the ideal reference, inulin. Inulin-based FE was 98.7% for D-asparagine and 84.8% for D-serine (28, 29). The initial D-asparagine-based FE of D-alanine (FE D-Ala/D-Asn) was 14.0 ± 5.8% (Figure 3, Table 3, Supplementary Figure S5). Just after oral intake of D-alanine at the low dose, FE D-Ala/D-Asn decreased because of the rapid increase in the blood D-alanine level. After this, FE D-Ala/D-Asn increased quickly to its peak of 64.5 ± 10.3%, followed by a decline and became constant at about 30%. Suppose the glomerular filtration rate, and its almost identical D-asparagine clearance, was 120 mL/min; the urinary clearance of D-alanine at the baseline and the peak and constant stages was estimated to be 16.8, 77.4, and 36.0 mL/min, respectively. It was suggested that the urinary excretion of D-alanine increases when the blood level increases. The T_*max*_ was 1.90 ± 0.56 h. The initial decline of the FE likely represented the time-lag between the peak concentration of D-alanine in plasma and urine. The FE of L-alanine, on the other hand, was constant throughout the study (Supplementary Figure S5). The dynamics of FE D-Ala/D-Asn was close to D-serine-based FE D-alanine (FE D-Ala/D-Ser, Figure 3 and Supplementary Figure S5). The analysis of the high-dose data showed a similar trend, with the C_*max*_ of FE D-Ala/D-Asn being 87.3 ± 4.3% (Figure 3 and Table 3).

**Table 3.** Kinetics of fractional excretion of D-alanine.

| **Dose** | **C<sub>0</sub>** |  |  | **T<sub>max</sub>** |  |  | **C<sub>max</sub>** |  |  |
| --- | --- | --- | --- | --- | --- | --- | --- | --- | --- |
| **(μmol)** | **(%)** |  |  | **(h)** |  |  | **(%)** |  |  |
| 11,236 | 14.0 | ± | 5.8 | 1.90 | ± | 0.56 | 64.5 | ± | 10.3 |
| 33,708 | 25.4 | ± | 5.1 | 1.70 | ± | 0.12 | 87.3 | ± | 4.3 |

%, fractional excretion of D-alanine ratio with D-asparagine as a reference.

**Figure 3.**
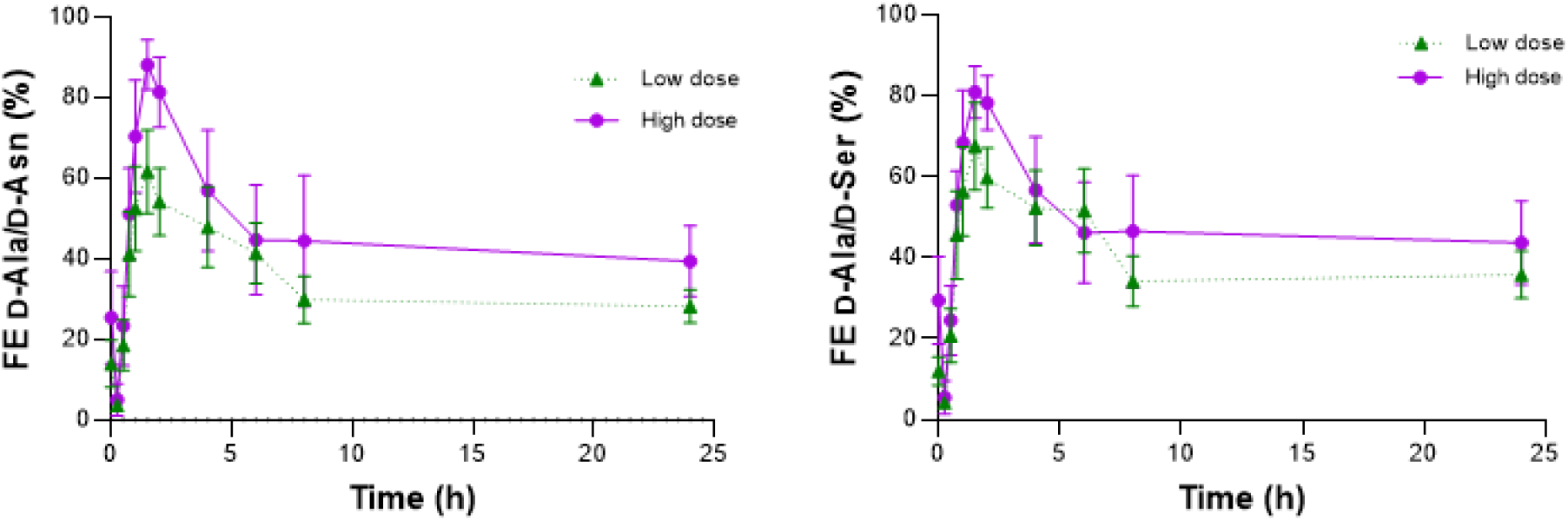
Fractional excretion of D-alanine after oral intake of D-alanine. Fractional excretion (FE) was calculated using D-asparagine or D-serine as references.

### Analysis of other D-amino acids

We monitored the changes of other amino acids after oral intake of D-alanine. For this purpose, we selected asparagine, serine, and proline, whose D-forms are relatively abundant in blood (3, 7). After oral intake of D-alanine, the levels of D-asparagine and D-serine were largely unchanged (Supplementary Figure S1 and S2). The level of D-proline showed a declining trend; however, this association was ambiguous because of the wide error bars. Similarly, the changing trends of plasma levels of D-amino acids were unclear in the analysis of the high-dose data. The levels of L-amino acids were largely unchanged, probably because of their larger volumes in the body. The level of L-asparagine might have shown a transient increase after oral uptake of D-alanine at the high dose. This change was only transient and decreased back to baseline. Urinary ratios of D-asparagine and D-serine were largely unchanged (Supplementary Figure S3 and S4).

## Discussion

This study unraveled the dynamics of D-alanine in the body after oral intake. Upon oral intake, D-alanine is quickly absorbed and enters the bloodstream. The plasma level of D-alanine peaks about 30 min after oral intake, followed by exponentially quick clearance. The blood level reaches close to the endogenous level after 24 h. The D-alanine that enters the bloodstream is delivered to the kidney and excreted into urine. The peak in urinary level follows the blood level and is reached about 1 h after oral intake, reflecting the time lag between it entering the blood and urinary excretion. The FE of D-alanine increases upon oral intake, and the significance of urinary clearance as part of whole-body clearance was assessed. This provides key insight into the physiology of D-alanine.

D-Alanine is efficiently absorbed like L-alanine in the digestive tract. Orally ingested D-alanine quickly appears in blood, and the blood level of D-alanine shows a dose-dependent increase to the same range as L-alanine. D-Alanine is present in food, and eating foods rich in D-alanine can induce an increase in the blood D-alanine level (18). The amino acid transporters are responsible for the absorption of D-alanine. Transporters recognize the chirality of the amino acid for chiral-selective transportation (4, 7). The efficacy of the D-alanine transport system is likely high. Many amino acid transporters remain uncharacterized, especially for D-amino acids, usually found in trace amounts (33). In the case of D-serine, a relatively well-studied D-amino acid, only five transporters are known to deliver D-serine (4, 33); however, a whole body of transporter system is far from elucidation. Currently-uncharacterized transporters are thought to deliver D-alanine.

The volume of distribution of D-alanine was estimated to be about 9 L, regardless of the dose. This volume exceeds the plasma volume of ∼ 4.6 L, if the body weight is assumed to be 60 kg. Therefore, D-alanine is distributed outside the plasma and delivered to the tissues. Additionally, the distribution volume is likely much less than that of L-alanine. Since the tissue distribution is limited and FE is high, D-alanine in the blood is efficiently excreted into urine. Within the estimated 200 mL/min clearance, urinary excretion is responsible for the 16.8−77.4 mL/min. In the case of L-alanine, FE is close to 1%, suggesting almost negligible urinary excretion. Despite this, the level of L-alanine in the blood decreases quickly from the peak and becomes constant after oral uptake. This fact suggests that the distribution volume of L-alanine is higher than that of D-alanine, and orally ingested L-alanine is quickly re-delivered to the tissues.

In response to oral uptake, the urinary clearance of D-alanine increases. After glomerular filtration, D-alanine is inefficiently reabsorbed by the proximal tubules, while L-alanine is almost completely reabsorbed. This is reflected in the higher FE of D-alanine compared to that of L-alanine. In an adult with a GFR of 120 mL/min, 77.4 mL/min of D-alanine in the blood is cleared from the urine at peak, while the proximal tubules absorb the rest (42.6 mL/min). The transporter system for D-alanine is inefficient in the proximal tubules (4). Despite this, the FE of D-alanine did not reach 100% when the blood level of D-alanine increased, and the original urine just after glomerular filtration was assumed to contain much higher level of D-alanine. Instead of reaching its maximum capacity to reabsorb D-alanine, the proximal tubules’ transporter system can upregulate the amount reabsorbed.

The reabsorbed portion of D-alanine in the proximal tubule either re-enters the bloodstream or is oxidized by DAO. While DAO is predominantly present in the proximal tubules of the kidney, it is also present in brain, spinal cord, liver, neutrophils, retina, and small intestine (24, 34). Based on our estimation, DAO plays a major role in the clearance of D-alanine. Among the reabsorbed portion of D-alanine, at least a portion (X %) of reabsorbed D-alanine is directly oxidized by DAO. Suppose that no other enzyme is responsible for the metabolism of D-alanine; the potential amount of D-alanine subject to oxidation by DAO in the body after the oral intake is estimated as follows:

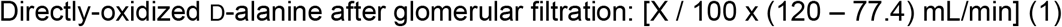

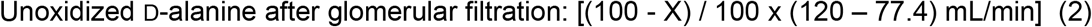

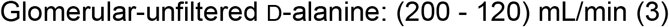

The sum of (2) and (3), (122.6 – 42.6 x X / 100) mL/min, is the potential target of DAO oxidation. This D-alanine portion is oxidized outside the kidney or in the proximal tubule through basolateral entry. D-Alanine potentially enters the proximal tubule from the basolateral side without glomerular filtration. The basolateral entry of amino acids is usually small and negligible in contrast to the massive amount of reabsorption from the original urine, whereas it increases when the levels of amino acids increase. In metabolic acidosis, for example, the blood level of glutamine increases because of release from muscle. Then, the kidney proximal tubule takes up glutamine from both the tubular lumen and the basolateral side (35). Likewise, DAO in the proximal tubule may also play a role in the clearance of glomerular-unfiltered D-alanine. The plasma volume of the kidney artery is about 500 mL/min, meaning the DAO in the kidney can fully cover the metabolism of the glomerular-unfiltered D-alanine. DAO in the kidney and extra-kidney cooperate in the clearance of D-alanine.

This study indirectly estimated the contribution of DAO in the clearance of D-alanine. This may be in keeping with the blood levels of D-alanine in patients with kidney disease or *Dao*-deficient rodent. The blood level of D-alanine increases up to 82 μmoL in patients with kidney disease (3). The increase in the blood D-alanine level in kidney failure is a result of (i) reduced kidney blood flow and glomerular filtration and subsequent urinary excretion, (ii) reduced delivery of D-alanine from the basolateral side to the proximal tubule, and (iii) reduced oxidation in the kidney. Among them, the reduced kidney blood flow is the most upstream factor that induces an increase in D-alanine level. In mice lacking DAO activity, the blood level of D-alanine was elevated to about 50 μM with a striking increase in urinary level (26). Under *Dao*-deficient conditions, basolaterally delivered D-alanine is not oxidized in the kidney, and extra-kidney organs also lack the capacity to oxidize D-alanine. Thus, the blood D-alanine level is kept high, leading to the high urinary excretion.

Orally-ingested D-alanine is essential for homeostasis. During severe viral infection, the level of D-alanine decreases (5, 6). While D-alanine mediates signal transduction in cells (12), D-alanine also exerts its physiological function through oxidation (9, 12). Therefore, a reduced level of D-alanine suggests an increase in the oxidation of D-alanine. Supplementation of D-alanine is protective against viral infections in model mice. Mechanistically, D-alanine regulates the immunological response directly or indirectly through regulating the circadian rhythm (9, 12). D-Alanine can normalize the circadian rhythm. Additionally, D-alanine induces the expression of circadian clock genes, regulating of a board range of circadian-related biological processes, such as metabolic processes and longevity-regulating pathways. With its intrinsic circadian rhythm, D-alanine maintains whole sets of physiological functions in the body. D-Amino acids can directly affect the function of the kidney. Orally-ingested D-alanine likely modifies the transporter activity of the kidney. As seen with D-serine, there might be a chiral feed-back system for D-alanine.

This study had several limitations. The number of participants was limited. The clearance of D-alanine by DAO was indirectly estimated based on the concept that D-alanine is not cleared through other biological processes. Despite this, the precise measurement of D-alanine potentiates a detailed kinetic analysis.

In conclusion, we quantified the significance of kidney urinary clearance and the remnant clearance by DAO. While D-alanine is efficiently absorbed after oral intake, D-alanine is delivered to tissues with a distribution volume larger than the plasma volume. Urinary excretion and DAO cooperate in maintaining D-alanine levels in the body. This kinetic analysis adds to the basic knowledge of the modulation of physiological processes after the oral intake of D-alanine.

## Data Availability

All data produced in the present work are contained in the manuscript.

